# Stress and Coping Strategies among Medical Students in Dubai, United Arab Emirates, in 2020: A Cross-Sectional Study

**DOI:** 10.1101/2022.09.24.22280306

**Authors:** Yas Kaveh Boushehri, Lakshmanan Jeyaseelan, Meshal A. Sultan

## Abstract

Studies across the world, have revealed elevated levels of stress among medical students. The rate of significant stress is 55% higher among healthcare professionals in comparison to the general population. This level of stress may lead to higher rates of burnout, depression, and functional impairment. This study aims to investigate the stress levels among medical students in Dubai and also to assess their coping strategies. The total number of participants in this study was 97. Rates of high stress levels as per the Perceived Stress Scale (PSS-10) was found to be higher among year 1 to year 3 medical students (43.6%), in comparison to year 4 and year 5 medical students (7.7%). The Brief-COPE inventory was used in this study and found that among medical students the mean score for a Problem Focused coping style was 22 out of 32 (medium to high range). Future research that evaluates a more comprehensive investigation into the psychological impact of stress and also exploration of effective strategies to enhance coping with stress is highly warranted.

## INTRODUCTION

Elevated stress levels have been reported among medical students and also in the general population. Additionally, ongoing high levels of stress have been found to have a significant negative impact on an individual’s ability to function. Studies have been conducted around the world to explore any possible relationship between the onset of the COVID-19 pandemic and the prevalence of distress.

A large-scale general population survey of psychological distress was conducted in China between January 31st to February 10th, 2020 (Qiu et al., 2020). Of the 52730 subjects studied, 5.1% reported severe distress levels (Qiu et al., 2020). Another study using questionnaires completed by the general public was conducted in China between January 31st and February 2nd, 2020. This study revealed severe stress levels among 2.6% of the 1120 respondents (Wang et al., 2020). A cross sectional survey in April 2020 that included 354 participants from the general population in India, reported 11.6% of respondents had moderate to severe stress levels (Verma and Mishra, 2020). Furthermore, a survey used in a cross sectional study among 1597 individuals from the general population in Saudi Arabia in May 2020, has reported severe stress levels among 6.4% of the participants (Alamri et al., 2020).

According to existing literature, high levels of perceived psychological stress and depression have been identified among medical students in Egypt, Saudi Arabia and United Arab Emirates (Elzubeir et al., 2010). A study that explored levels of stress among 938 postgraduate physicians enrolled in residency training programs in various medical specialties in Saudi Arabia revealed high levels of stress among 32.4% of the participants (Alosaimi et al., 2015). Additionally, a survey among year 1 to year 5 medical students in Saudi Arabia 2018 has shown high stress level among 51.7% of the study population (Shadid et al., 2020). Another study among medical students in Saudi Arabia conducted in 2016 reported high stress level among 16.5% of the participants (Siddiqui et al., 2017). A study among first year medical students in four universities in Malaysia conducted during the academic year 2008/2009 revealed that the prevalence of distress was approximately 50% (Yussof et al., 2011). The top five coping strategies among the participants in this study were religion, active coping, positive reinterpretation, acceptance, and planning (Yussof et al., 2011).

Elevated stress can have a negative impact on academic performance (Sohail, 2013; Elias et al., 2011; Melaku et al., 2015), as well as physical and psychological wellbeing (Abdulghani et al., 2011; Guthrie et al., 1998). Moreover, studies have shown that such high levels of stress can have negative effects on the cognitive functioning of medical students, impacting their ability to learn (Sohail, 2013; Elias et al., 2011; Melaku et al., 2015). Studies have revealed that chronic stress can lead to increased anxiety levels. (Rahman et al., 2016). Furthermore, structural brain changes have been reported as a result of chronic stress. For example a decline in hippocampal volume, which subsequently negatively impacts cognitive functioning has been reported (Wang et al., 2010; Rahman et al., 2016; Humphreys et al., 2017; Koskinen et al., 2020).

Therefore, it is imperative to evaluate stress levels among medical students in the hope of tackling this important issue and preventing further negative consequences. Additionally, effective stress management strategies can contribute to maximizing medical students’ learning experience. This study explores stress levels among medical students, how they adjust to their challenges, and how this correlates to non-medical participants.

## METHODS

### Participants and Procedures

The design of the study was a cross-sectional study and is based on the STROBE reporting guidelines (Cuschieri, 2019). Data collection involved online surveys based on standardised rating scales screening for psychological distress/ morbidity. The instruments used in this study were the Perceived Stress Scale (PSS-10) and the Brief COPE inventory. The online survey was sent via email to all medical students enrolled at a local university in the UAE, as well as to a group of selected non-medical individual’s. The online surveys were completed by the study participants between November 10th, 2020 and November 23rd, 2020.

The participants of this study were medical students from a local university and individuals recruited from outside the medical field. For medical student recruitment, all students enrolled at the local university during the academic year 2020/2021 were invited to participate in the study. For the non-medical participants, invitations were sent by the principal investigator to a personal friend group of individuals aged between 16 and 30 years old. This age range was chosen as all of the year 1-5 medical students in the study were also within this age range.

Participants in the study include nationals from the following countries Austria, Canada, Egypt, Germany, Great Britain, Jordan, India, Iran, Iraq, Italy, Kenya, Norway, Pakistan, Palestine, Saudi Arabia, South Africa, South Korea, Syria, UAE, USA, and Yemen. The nationality of participants were categorised as: 1. Gulf Cooperation Council (GCC) and Asian Countries, 2. United Arab Emirates (UAE), and 3. Europe, Canada, USA, South Africa, and others.

### Measurements

Data collection involved demographic variables such as gender, age, nationality, and year of study; as well as two rating scales to assess stress levels and coping strategies. The Perceived Stress Scale (PSS-10) implemented in this study assessed an individual’s level of stress during the previous month (Cohen et al., 1983; Cohen and Williamson, 1988). The questions evaluated in the PSS-10 were 1. How often have you been upset because of something that happened unexpectedly? 2. How often have you felt that you were unable to control the important things in your life? 3. How often have you felt nervous and stressed? 4. How often have you felt confident about your ability to handle your personal problems? 5. How often have you felt that things were going your way? 6. How often have you found that you could not cope with all the things that you had to do? 7. How often have you been able to control irritations in your life? 8. How often have you felt that you were on top of things? 9. How often have you been angered because of things that happened that were outside of your control? 10. How often have you felt difficulties were piling up so high that you could not overcome them? The PSS-10 scores range from 0 to 40 and are divided as per the following: scores ranging from 0-13 are considered low stress; scores ranging from 14-26 are considered moderate stress; scores ranging from 27-40 are considered high perceived stress.

Furthermore, the Brief COPE inventory assessed positive and negative coping strategies adopted at times of stress (Heffer and Willoughby, 2017). The same surveys were sent to individuals from the personal friend group and to the medical students.

The Brief COPE is a 28-item questionnaire used in the identification of 14 theoretical coping responses among individuals, including Self-distraction (items 1 and 19), Active coping (items 2 and 7), Denial (items 3 and 8), Substance use (items 4 and 11), Use of emotional support (items 5 and 15), Use of instrumental support (items 10 and 23), Behavioural disengagement (items 6 and 16), Venting (items 9 and 21), Positive reframing (items 12 and 17), Planning (items 14 and 25), Humour (items 18 and 28), Acceptance (items 20 and 24), Religion (items 22 and 27), and Self-blame (items 13 and 26) (Carver, 1997; Monzani et al., 2015). The coping styles were divided into the following three categories during the analysis: Problem-Focused, Emotion-Focused, and Avoidant Coping by combining related coping responses (Carver, 1997; Poulus et al., 2020). Problem-Focused coping included a total of eight items 2, 7, 10, 12, 14, 17, 23, and 25 as per the questions listed by Carver, 1997. The total score range for Problem-Focused coping is from 8 to 32. The range of total low score for this coping style is from 8 to 16 and the range of total medium to high score is from 17 to 32. Emotion-Focused coping included a total of twelve items 5, 9, 13, 15, 18, 20, 21, 22, 24, 26, 27 and 28 as per the questions listed by Carver, 1997. The total score range for Emotion-Focused coping is from 12 to 48. The range of total low score for this coping style is from 12 to 24 and the range of total medium to high score is from 25 to 48. Avoidant coping included a total of eight items 1, 3, 4, 6, 8, 11, 16, and 19 as per the questions listed by Carver, 1997. The total score range for Avoidant coping is from 8 to 32. The range of total low score for this coping style is from 8 to 16 and the range of total medium to high score is from 17 to 32.

High scores of Problem-Focused coping indicates strategies that aim to change the stressful situation and reflect psychological strength (Carver, 1997; Poulus et al., 2020). Emotion-Focused coping reflects strategies that aim to regulate emotions associated with the stressor (Carver, 1997; Poulus et al., 2020). High or low scores are not necessarily associated with positive or negative psychological health, however, they describe the individual’s coping style (Carver, 1997; Poulus et al., 2020). Regarding an Avoidant coping style, high scores represent efforts to disengage, while low scores represent healthy adaptive coping (Carver, 1997; Poulus et al., 2020).

An additional quantitative variable assessed in this research study was a ‘general level of stress’ on a scale of 1-10, 1 being the lowest level of stress and 10 being highest level of stress.

### Statistical Analysis

The sample size calculation was based on the prevalence estimate. The prevalence of severe stress was reported to be 52% by Shadid et al (2020). In order to estimate this with the precision of 10% with 95% CI, we need to study 96 medical students from years 1 to 5.

The prevalence of severe stress was presented according to both Medical and Non-Medical categories. This was presented according to socio demographic variables. Besides the prevalence of severe stress, the association between severe stress and categorical socio demographic variables such as nationality etc. were analyzed using chi-square test. The mean age between the severe and low or moderate stress was done using t test. The correlation between age and stress score estimated used Pearson correlation coefficient. The multivariable logistic regression analysis was done using logit link function with Severe stress vs Mild or moderate stress as outcome.

In Coping, we have calculated the summative for score for each domain such as Problem-Focused coping etc. In order to compare the means of the domain score according to socio demographic categorical variables such as gender, nationality etc., t and Analyses of variance (ANOVA) tests were done. We have presented the actual p values as this is more informative. Data was analyzed using SPSS Version 24.

### Ethical Approval and Informed Consent

No individuals were excluded from this study based on racial, gender, religious or cultural backgrounds. Consent was obtained from participants age 18 years and above. Assent was obtained from participants below age 18 years along with consent from their parents. This study was conducted with the approval of the local ethical review board.

## RESULTS

A total of 235 medical students from year 1 to year 5 were invited to participate in the study. This number represents all medical students enrolled during the academic year 2020/2021 as per the following: 53 in Year 1, 52 in Year 2, 51 in Year 3, 35 in Year 4, and 44 in Year 5. The number of medical students who participated was 87. The total number of non-medical individuals who were invited to the study was 10. The number of non-medical individuals who participated was 10. This represents a 41.3% response rate (Figure 1).

**Figure 1.**
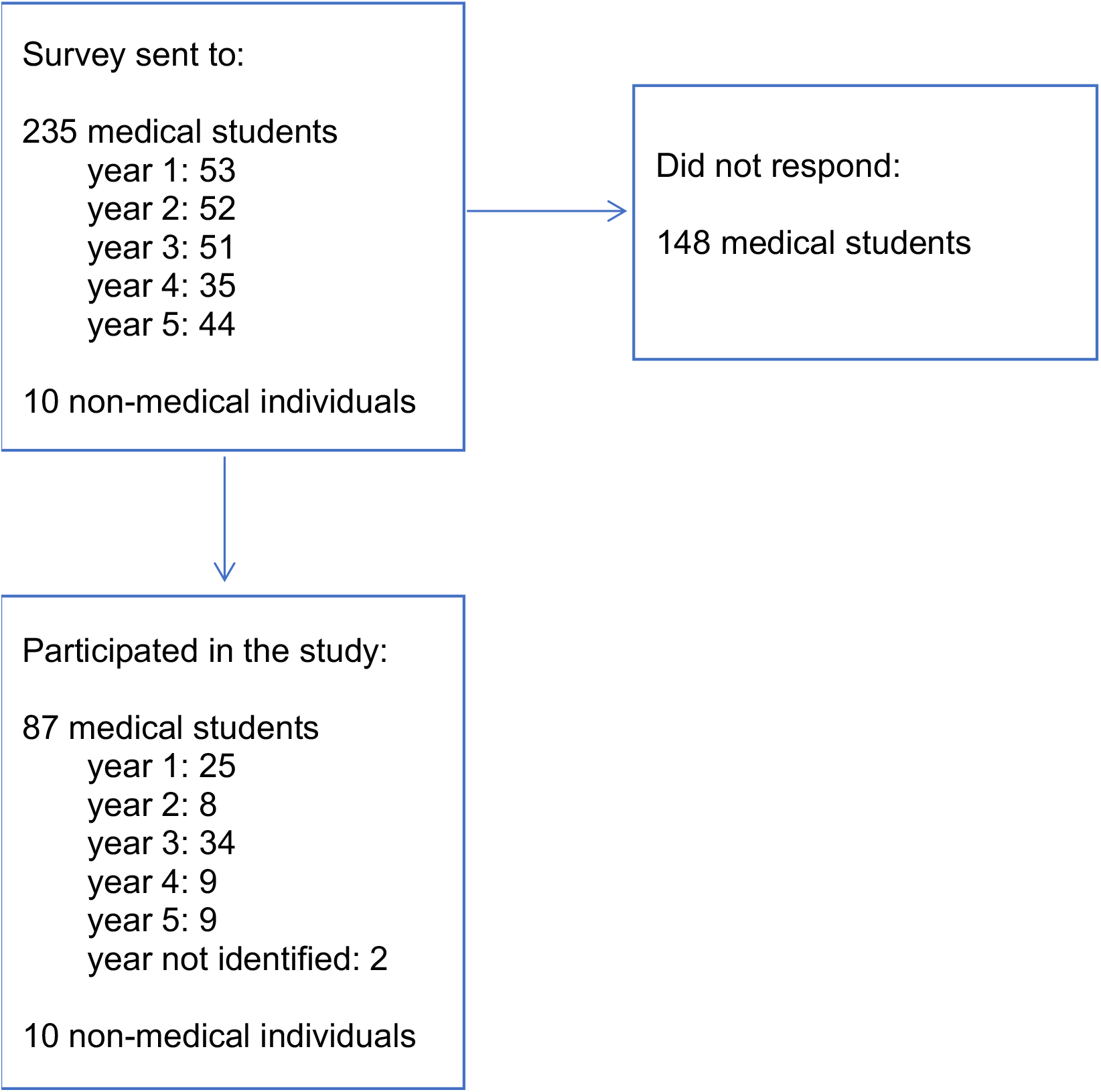
Flow Chart of Participants

It was found that the majority of participants were female. The gender distribution was 25% males and 75% females in medical and 40% male and 60% female in the non-medical category. About 32% of medical students are from UAE, while this was 20% in non-medical participants. In the medical category 45% are from Europe, Canada, USA South Africa and others, while this was 70% in the non-medical participants. GCC and Asian countries participants are 23% and 10% in the medical and non-medical groups respectively. About 38% of medical students are from first and second years. About 39% and 21% are from year 3 and year (4 or 5) respectively. The difference in the distribution was significant (p=.001). The mean (sd) age was 20 (2.3) in medical category which was significantly lower when compared to 24 (2.4) years in non-medical category (p=.001).

After asking both groups of participants to indicate their general level of stress on a scale of 1-10, the mean (SD) stress levels obtained for medical and non-medical participants were 7 (2) and 6 (2), respectively.

The Perceived Stress Scale (PSS-10) numerical scores obtained for medical and non-medical participants were then used to identify the corresponding stress levels for individuals. The PSS-10 scores obtained demonstrated that the majority of medical and non-medical participants experienced moderate stress levels. However, high perceived stress was reported at a higher rate among medical participants 42.5% (95% CI: 32.6, 52.3) than non-medical participants 20% (95% CL: 12.0,27.9). Furthermore, according to the PSS-10, only 5.8% of medical students experienced low stress while among non-medical participants, 30% experienced low stress. Table 1 below shows the number and percentage of medical and non-medical participants, respectively, for each PSS-10 score.

**Table 1.**
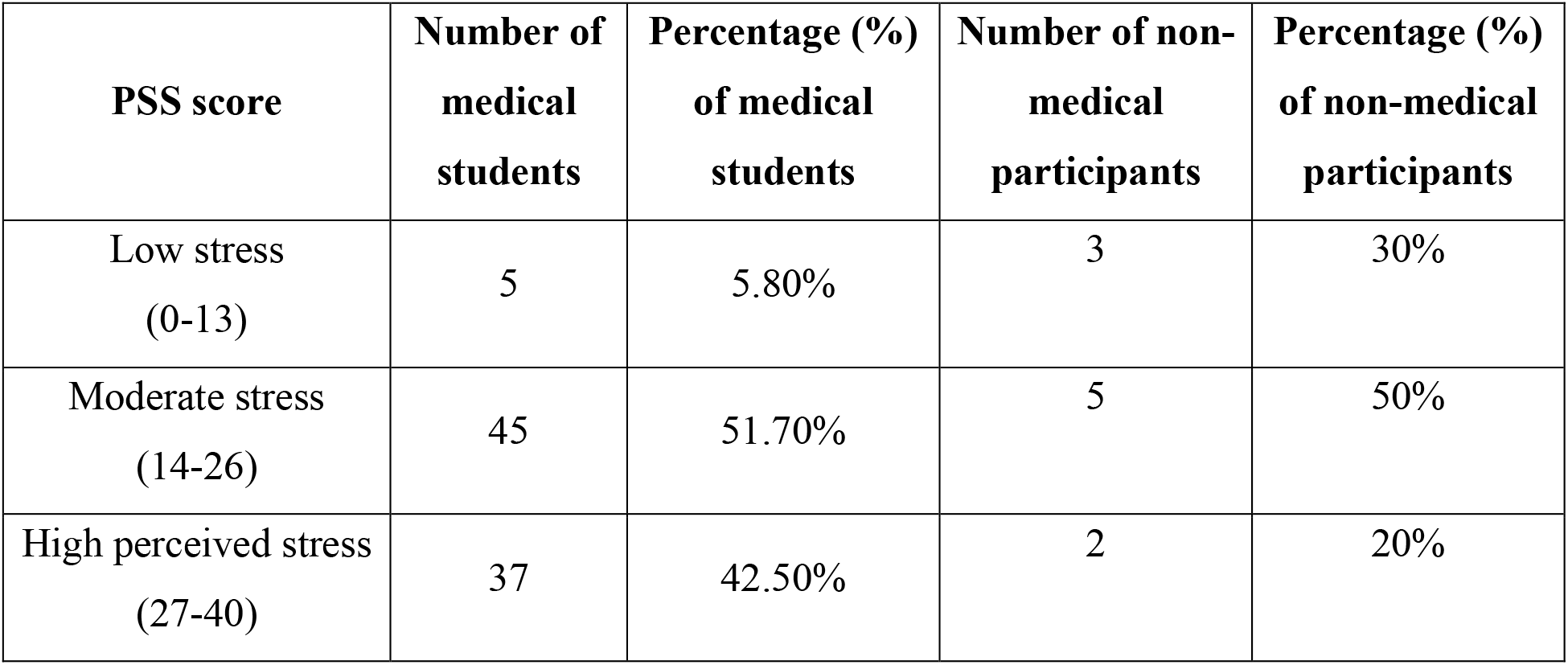
The number and percentage of medical students and non-medical participants allocated to each PSS-10 score (n=97)

| **PSS score** | **Number of medical students** | **Percentage (%) of medical students** | **Number of non-medical participants** | **Percentage (%) of non-medical participants** |
| --- | --- | --- | --- | --- |
| Low stress<br>(0-13) | 5 | 5.80% | 3 | 30% |
| Moderate stress<br>(14-26) | 45 | 51.70% | 5 | 50% |
| High perceived stress<br>(27-40) | 37 | 42.50% | 2 | 20% |

The Brief COPE inventory gathered data in relation to the common coping mechanisms adopted by medical and non-medical participants. According to the findings, the 3 most common coping responses adopted by medical students were self-distraction, humour, and planning, while non-medical participants tend to commonly adopt active coping, self-blame, and positive reframing. Moreover, denial, behavioral disengagement and substance use were the least common coping responses among medical students. In contrast, venting, denial, and substance use were not demonstrated among any of the non-medical participants. Graphs 1 and 2 below demonstrate the results obtained using the Brief COPE inventory.

**Graph 1.**
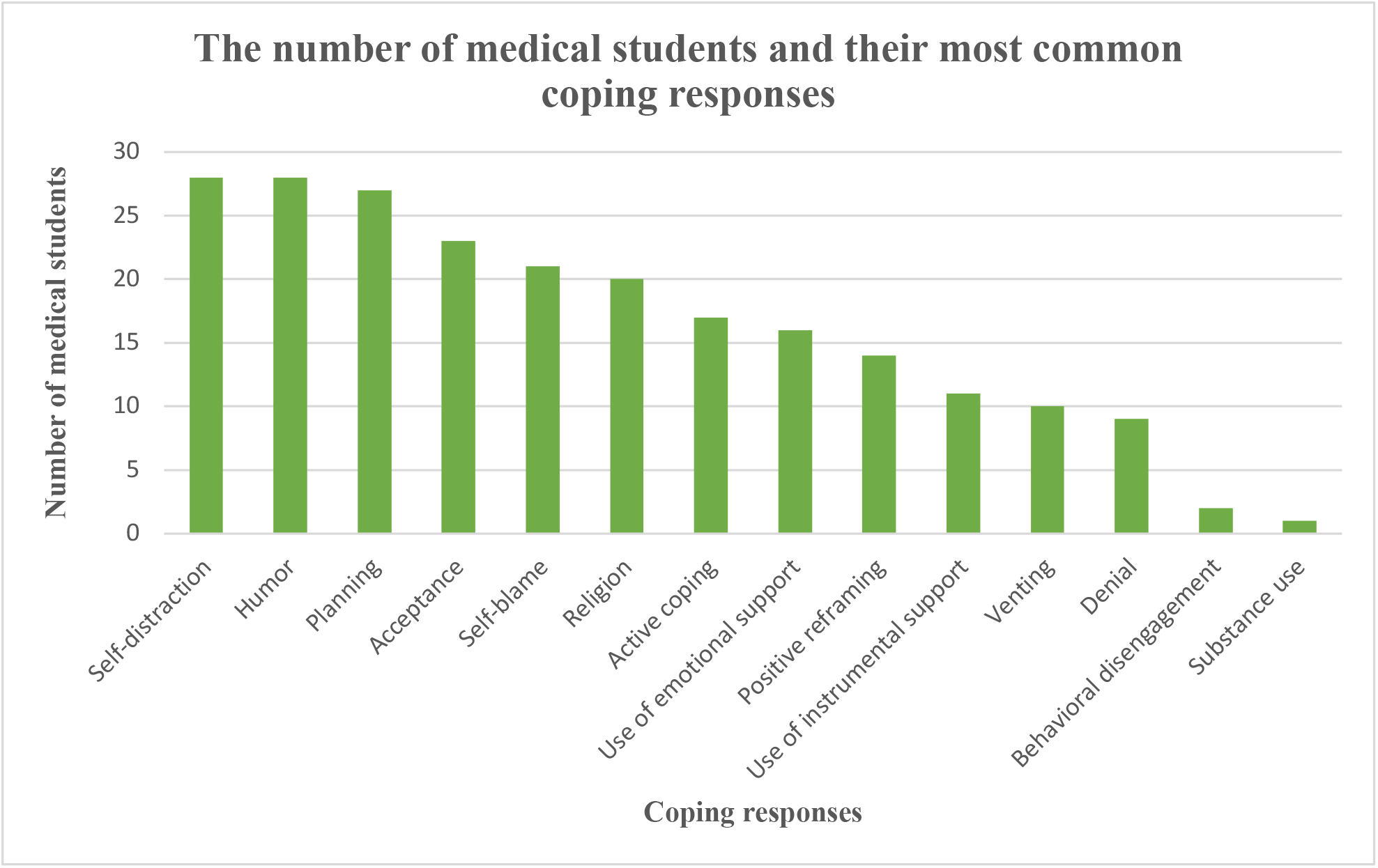
The number of medical participants and their most common coping responses, according to the brief COPE (n=87)

**Graph 2.**
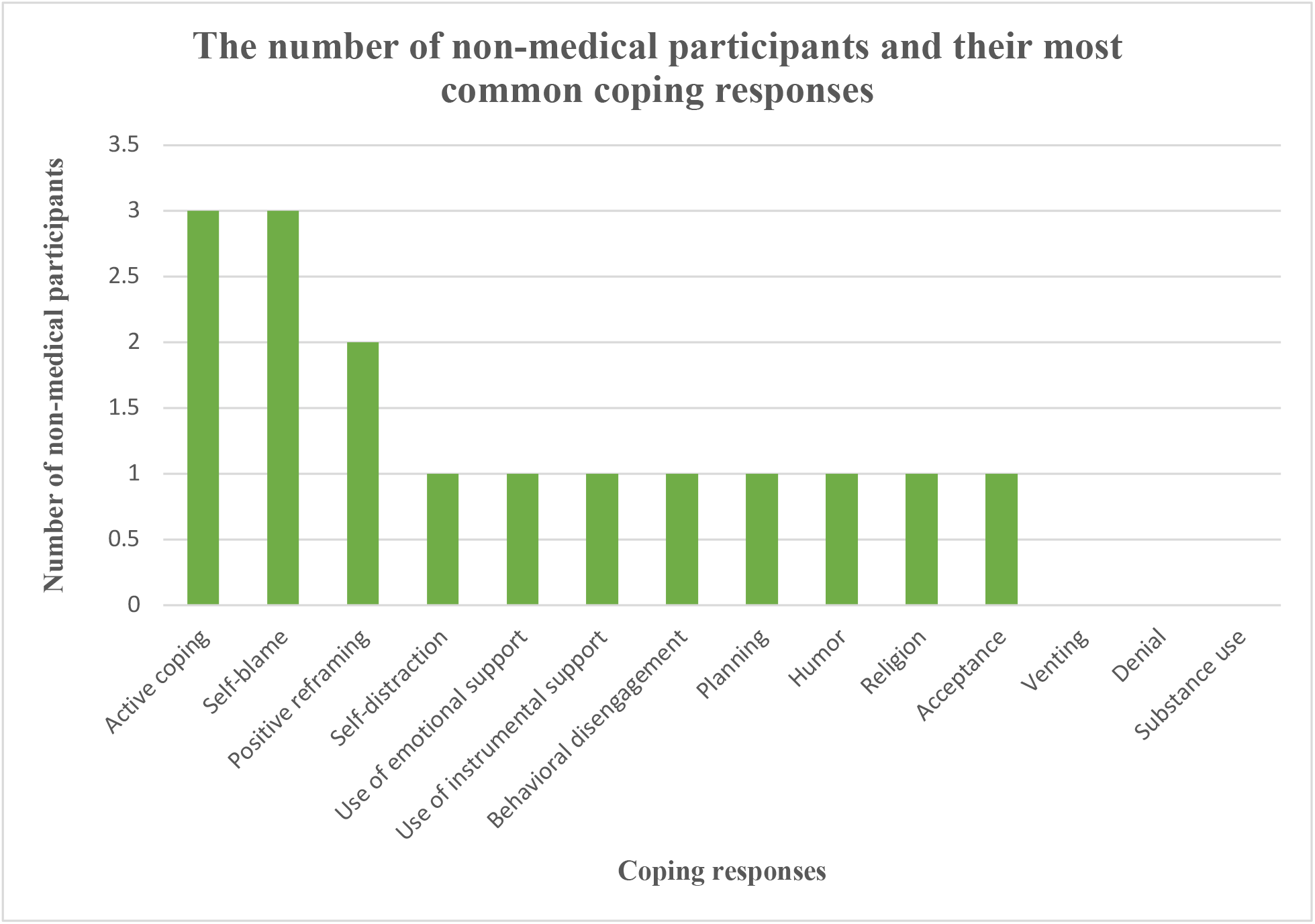
The number of non-medical participants and their most common coping responses, according to the brief COPE (n=10)

Table 2 shows the mean of the general levels of stress on a scale of 1-10 as well as the PSS-10 scores. Year 3 medical students obtained the highest mean general level of stress (7.53/10), followed by year 5 (7.44/10), year 1 (7.04/10), year 2 (7/10) and year 4 (6/10) having the lowest mean general level of stress. Year 3 medical students obtained the highest mean PSS-10 score of (25.79), followed by year 1 (25.56), year 2 (23.00), year 5 (22.22), and year 4 (20.33) having the lowest mean PSS score. However, despite these differences in PSS-10 scores, the corresponding stress levels for all 5 years is moderate stress as it is within the range of 14-26.

**Table 2.** The mean general levels of stress (1-10), mean numerical PSS-10 scores, and corresponding stress levels for year 1-5 medical students (n=85)

| **Number per year of study** | **General level of stress mean (1-10)** | **PSS-10 score mean (numerical)** | **PSS-10 (stress level)** |
| --- | --- | --- | --- |
| Year 1 (n=25) | 7.04 | 25.56 | Moderate |
| Year 2 (n=8) | 7 | 23 | Moderate |
| Year 3 (n=34) | 7.53 | 25.79 | Moderate |
| Year 4 (n=9) | 6 | 20.33 | Moderate |
| Year 5 (n=9) | 7.44 | 22.22 | Moderate |

Analysis of the socio-demographic variables and stress level revealed a statistically significant difference (p=.018) in the association between stress level and age (Table 3). The mean (sd) for age of participants reporting high stress scores on PSS-10 was 19.5 years (2.3). The mean (sd) for age of participants reporting low to moderate stress scores on PSS-10 was 20.8 years (2.6). Among the total of 39 participants who reported a high level of stress as per PSS-10 scores, 43.6% were Year 1 or 2 medical students, 43.6% were Year 3 medical students, 7.7% were Year 4 or 5 medical students, and 5.1% were non-medical participants (p=.028). Among the total of 58 participants who reported low to moderate levels of stress as per PSS-10 scores, 29.3% were Year 1 or 2 medical students, 29.3% were Year 3 medical students, 27.6% were Year 4 or 5 medical students, and 13.8% were non-medical participants (p=.028).

**Table 3:**
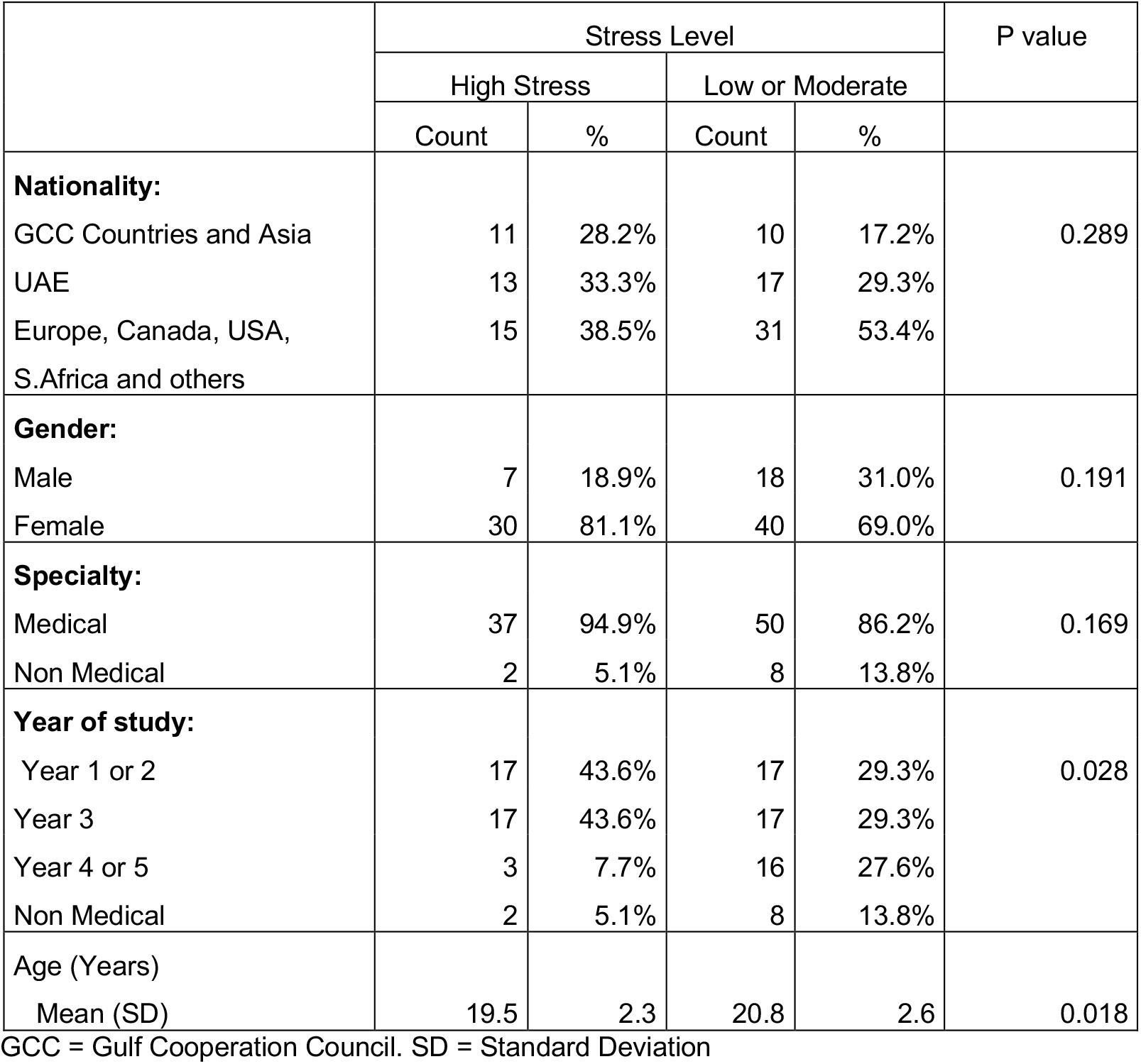
Association between Socio Demographic variables and Stress Level

|  | Stress Level |  |  |  | P value |
| --- | --- | --- | --- | --- | --- |
|  | High Stress |  | Low or Moderate |  |  |
|  | Count | % | Count | % |  |
| **Nationality:** |  |  |  |  |  |
| GCC Countries and Asia | 11 | 28.2% | 10 | 17.2% | 0.289 |
| UAE | 13 | 33.3% | 17 | 29.3% |  |
| Europe, Canada, USA,<br>S.Africa and others | 15 | 38.5% | 31 | 53.4% |  |
| **Gender:** |  |  |  |  |  |
| Male | 7 | 18.9% | 18 | 31.0% | 0.191 |
| Female | 30 | 81.1% | 40 | 69.0% |  |
| **Specialty:** |  |  |  |  |  |
| Medical | 37 | 94.9% | 50 | 86.2% | 0.169 |
| Non Medical | 2 | 5.1% | 8 | 13.8% |  |
| **Year of study:** |  |  |  |  |  |
| Year 1 or 2 | 17 | 43.6% | 17 | 29.3% | 0.028 |
| Year 3 | 17 | 43.6% | 17 | 29.3% |  |
| Year 4 or 5 | 3 | 7.7% | 16 | 27.6% |  |
| Non Medical | 2 | 5.1% | 8 | 13.8% |  |
| **Age (Years)** |  |  |  |  |  |
| Mean (SD) | 19.5 | 2.3 | 20.8 | 2.6 | 0.018 |

GCC = Gulf Cooperation Council. SD = Standard Deviation

Table 4 presents the findings of multivariable logistic regression analyses that was modelled for Severe stress. The female participants had 1.1 (95% CI:0.31, 3.8) times higher levels of severe stress as compared to male participants (p=.87). The GCC and Asian participants had 1.8 (95% CI:0.5, 6.6) times higher levels of severe stress as compared to the European and Canadian participants (p=.38). However, the UAE participants had about 15% less stress as compared to the European, Canadian and other participants from the various other countries (p=0.79). As the age of the participant increased by one year, the odds of getting severe stress was 30% less as compared to participants who were younger by one year (p=.15). The first- and third-year medical students had 1.5 and 2.0 times higher odds of getting severe stress as compared to 5^th^ year medical students, however, the association was not significant.

**Table 4:**
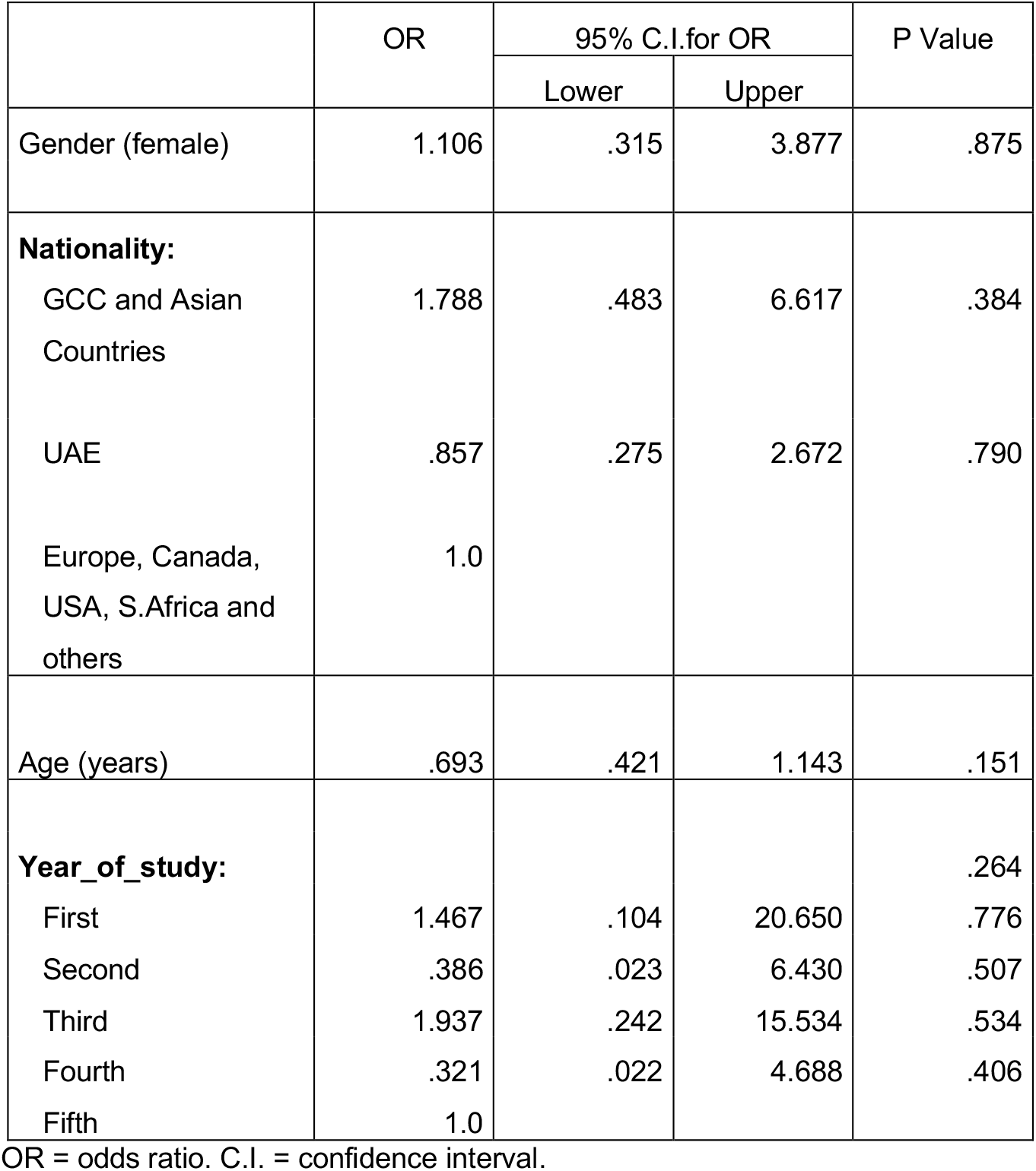
Multivariable Regression analyses: Risk factors for Severe Stress.

|  | OR | 95% C.I. for OR |  | P Value |
| --- | --- | --- | --- | --- |
|  |  | Lower | Upper |  |
| Gender (female) | 1.106 | .315 | 3.877 | .875 |
| **Nationality:** |  |  |  |  |
| GCC and Asian Countries | 1.788 | .483 | 6.617 | .384 |
| UAE | .857 | .275 | 2.672 | .790 |
| Europe, Canada, USA, S.Africa and others | 1.0 |  |  |  |
| Age (years) | .693 | .421 | 1.143 | .151 |
| **Year_of_study:** |  |  |  | .264 |
| First | 1.467 | .104 | 20.650 | .776 |
| Second | .386 | .023 | 6.430 | .507 |
| Third | 1.937 | .242 | 15.534 | .534 |
| Fourth | .321 | .022 | 4.688 | .406 |
| Fifth | 1.0 |  |  |  |

OR = odds ratio. C.I. = confidence interval.

### Coping

In regard to the analysis of coping styles, the overall mean (sd) of Problem Focused Coping, Emotion Focused Coping and Avoidant Coping was 21.3 (4.9), 31.1 (6.3), and 15.8 (3.5) respectively (Table 5). The mean (sd) score for Problem Focused Coping was 23.4 (5.1) among UAE nationals. This represents a moderate to high score for healthy coping strategies including active coping, use of instrumental support, positive reframing, and planning. Furthermore, the mean score among UAE nationals was higher as compared to other nationalities and the difference was statistically significant (p=.013). The mean (sd) score for Problem Focused Coping was 21.7 (4.9) among medical students and 18.3 (4.9) among non-medical participants. The difference in the scores between these two groups was statistically significant (p=.04). A statistically significant difference was also found between the scores of Emotion Focused Coping among medical students and non-medical participants (p=.008). The mean (sd) score for Emotion Focused Coping was 31.8 (6.2) among medical students and 26.2 (5.3) among non-medical participants. This represents more engagement in coping strategies that are based on use of emotional support, venting, self-blame, humor, acceptance, and religion. Moreover, a statistically significant difference (p=.028) was noted in the mean (sd) score of Emotion Focused Coping among Year 3 medical students 32.7 (5), in comparison to Year 1 or 2 medical students 31.8 (6.4), Year 4 or 5 medical students 29.7 (7.5), or non-medical participants 26.5 (5.7). In regards to Avoidant Coping style there was not any statically significant difference between participants as per nationality, gender, type of course, or year of study.

**Table 5:** Mean, SD of Coping strategies by Socio Demographic variables

|  | Problem Focused Coping |  | P Value | Emotion Focused Coping |  | P Value | Avoidant Coping |  | P Value |
| --- | --- | --- | --- | --- | --- | --- | --- | --- | --- |
|  | Mean | SD |  | Mean | SD |  | Mean | SD |  |
| **Nationality:** |  |  | **0.013** |  |  | **0.175** |  |  | **0.694** |
| GCC Countries & Asia | 19.62 | 4.66 |  | 31.57 | 5.53 |  | 16.43 | 4.34 |  |
| UAE | 23.40 | 5.08 |  | 32.73 | 6.69 |  | 15.60 | 3.44 |  |
| Europe, Canada, USA, S.Africa and others | 20.76 | 4.60 |  | 30.00 | 6.30 |  | 15.74 | 3.32 |  |
| **Gender:** |  |  | **0.178** |  |  | **0.127** |  |  | **0.808** |
| Male | 20.08 | 3.98 |  | 29.40 | 5.92 |  | 15.60 | 3.74 |  |
| Female | 21.63 | 5.18 |  | 31.66 | 6.40 |  | 15.80 | 3.45 |  |
| **Type of course:** |  |  | **0.040** |  |  | **0.008** |  |  | **0.960** |
| Medical | 21.68 | 4.85 |  | 31.76 | 6.21 |  | 15.84 | 3.35 |  |
| Non Medical | 18.30 | 4.85 |  | 26.20 | 5.25 |  | 15.90 | 5.36 |  |
| **Year of study:** |  |  | **0.442** |  |  | **0.028** |  |  | **0.93** |
| Year 1 or 2 | 21.94 | 5.23 |  | 31.82 | 6.38 |  | 15.85 | 3.48 |  |
| Year 3 | 21.56 | 4.99 |  | 32.74 | 4.99 |  | 15.74 | 3.18 |  |
| Year 4 or 5 | 21.00 | 4.08 |  | 29.74 | 7.53 |  | 15.74 | 3.80 |  |
| Non Medical | 19.10 | 5.26 |  | 26.50 | 5.74 |  | 16.40 | 5.02 |  |

SD = standard deviation

In terms of reliability for PSS-10, the overall Cronbach’s alpha was 0.874. In terms of reliability for Brief COPE, the overall Cronbach’s alpha was 0.813. These indicate a high degree of internal consistency of both tools.

## DISCUSSION

When assessing PSS-10 scores between medical and non-medical participants, it was found that both groups predominantly experienced moderate stress. With 51.7% and 50% respectively and with a higher rate of high perceived stress among medical students (42.5%) compared to non-medical participants (20%). According to a cross-sectional study comparing perceived stress among medical and non-medical students in Minya, Egypt, the prevalence of perceived stress was higher in medical students (88.9%) than in non-medical students (83.5%) (Seedhom et al., 2019). 18.8% of medical students experienced severe stress compared to 12.4% of non-medical students, which supports the results of this study indicating that medical students experienced higher levels of perceived stress than non-medical participants (Seedhom et al., 2019). Another cross-sectional study conducted in Riyadh, Saudi Arabia, explored perceived stress and associated factors among medical students. This study revealed a mean PSS score of 26.03, indicative of upper boundary moderate stress (Saeed et al., 2016). The study also discovered that severe stress was associated more with females and junior levels (Saeed et al., 2016). A study conducted in Jeddah, Saudi Arabia, assessed perceived stress and the reasons behind it among medical students. This study also obtained a high mean perceived stress score of 28.5, indicating that 52% of the participants were experiencing excessive stress (Gazzaz et al., 2018).

Our study showed mean age of participants reporting high stress scores was 19.5 years, which is lower than the mean age of participants reporting low to moderate stress levels which was 20.8 years (p=.018). Similarly, a previous study among medical students in Saudi Arabia revealed increased prevalence of high levels of stress among age 18 to 24 years (52.7%) in comparison to age 25 and above (37.5%) (Shadid et al., 2020). The analysis of reported high stress level and year of study in our sample revealed that year 3 medical students represented 43.6% of these responses (p=.028). This may be explained by the fact that year 3 is the final year before phase 3 of medical school clinical practice. Therefore, students experience greater stress due to the weightage of their courses and the additional course requirements in comparison to previous years. A study evaluating sources of stress among 152 medical students concluded that the main common stressors were academic-related and that it affected their performances in practicals, periodic examination performances and their general worrying about the future (Gazzaz et al., 2018). Another cross-sectional study among 359 medical students revealed that the top stressors were related to academics, for instance exams, large amount of contents to be learnt, getting poor marks, and lack of time to review what has been learnt (Yusoff et al., 2011).

The Brief COPE inventory highlighted the top 3 coping mechanisms among medical students as being self-distraction, humour and planning. Self-distraction tends to be a form of avoidance and attempting to run away from the problem rather than actually facing it. Therefore, this is not considered a healthy approach for medical students, taking into account the workload and necessity for time management, organization and self discipline. Coping responses can be further divided into approach and avoidance forms. Approach coping strategies facilitate ones ability to attain goals, whereas avoidance leads to temporary disengagement and abandonment of ones personal goals (Monzani et al., 2015). Humour tends to be both an avoidance and approach form of coping response. It may be said that when humour is identified in combination with other avoidance and unhealthy coping strategies, it can be considered as an avoidance strategy. However, when combined with other approach strategies such as planning, humour can be considered as a healthy coping response. According to a cross-sectional study assessing stress, coping, and burnout among final year-medical students, individuals with lower perceived stress scores more commonly adopted coping mechanisms such as acceptance, positive reframing, humour, planning and active coping (Singh et al., 2016).

In contrast, the top 3 coping responses for non-medical participants included active coping, self-blame and positive reframing. Active coping and positive reframing are considered as approach forms of coping, as opposed to self-blame which is identified as avoidance. Self-blame is considered an unhealthy form of coping strategy as it involves bringing oneself down rather than gaining the confidence and strength to push through and tackle problems (Monzani et al., 2015).

Furthermore, analysis revealed a mean (sd) score for Problem Focused Coping of 21.7 (4.9) among medical students and 18.3 (4.9) among non-medical participants (p=.04). This medium to high score reflects a tendency to change the stressor, plan the best course of action, suppression of competing activities, and seeking social support (Ogama, 2020). A previous study revealed that Problem Focused coping is associated with lower rates of burnout among medical students (Ogama, 2020). This was likely attributed to the impact of this coping style on lowering emotional exhaustion and increasing the sense of personal accomplishment (Ogama, 2020).

### Strengths and Limitations

This study is one of the first to explore stress levels and coping strategies among medical students in the UAE. One of the strengths is that participants have completed validated standardized rating scales. Furthermore, participants were from diverse nationalities. All 5 medical student cohorts were involved in data collection allowing for descriptive correlation of the year of study and stress levels. Moreover, the results of this research study have highlighted elevated stress levels among medical students relative to non-medical participants, encouraging further research with a larger sample size.

In terms of limitations, cross-sectional studies involve 1-time measurements of exposure and outcome and are therefore unlikely to produce accurate causal relationships (Xu et al., 2017). As the world is currently in the midst of a pandemic (Covid-19), stress levels assessed in this study may not be a result of the medical field but rather due to pandemic related behaviors such as self-isolation, altered sleeping patterns, lack of social activity, fearfulness, etc. Furthermore, participants may not be comfortable with certain questions and may conform to societal norms and beliefs, refraining from honest answers. Therefore, the assessment of substance use, for example, as a coping response may not be accurate. The majority of participants in this study were female and this could have had an impact on the results as, according to research, females tend to react differently to stressors, psychologically and biologically, as compared to males (Verma et al., 2011). The difference in stress levels between medical and non-medical participants may also be due to the difference in the mean ages of medical (23 years) and non-medical participants (19 years). Older participants may be less susceptible to stress potentially due to having a greater exposure to it, and having more experience dealing with stressors. Therefore, older participants may also be more acquainted with the management of stress. The small number of non-medical participants in the context of the availability of only 10 individuals from the friends’ group in the study age range may have not provided substantial data to accurately compare stress levels with that of medical students. We also did not collect information about current enrolment of non-medical participant in education. Therefore, we could not assess the relationships between stress related to medical school in comparison to other fields. Moreover, the PSS-10 assesses perceived stress over the last month and it therefore cannot provide a clear indication of chronic stressors and whether the individual is only experiencing high levels of perceived stress during that particular month or throughout the year. Therefore, the predictive validity of the PSS-10 score could decrease after a period of time due to changes in the exposure of stressors, life events, and coping resources available to the participants (Cohen and Williamson, 1988).

This study is prone to selection bias as the data population is clearly defined (Pannucci and Wilkins, 2010). Thus, data collected regarding stress levels and coping strategies may only be applicable to medical students in the local university, where the study was conducted, and not elsewhere. This could be prevented by collecting data from medical students in different universities. Confounding occurs when there is a third factor independently associated with stress levels, affecting establishment of an association between stress and the medical field, and thus, the study outcome (Pannucci and Wilkins, 2010). Such factors include the presence of a medical or mental disorder, which can result in significant stress levels. Moreover, students experiencing major life changes such as the death of a loved one or financial problems, could be experiencing immense levels of stress. Furthermore, the world could be in the midst of unforeseen circumstances, as is the case currently with regards to Covid-19 pandemic, resulting in substantial levels of stress worldwide. While observational studies cannot establish causal relationships, confounding factors should be taken into account when analysing stress levels in medical students and determining whether it is purely due to the medical field or external factors, or both. In formulating the survey for this study, the used rating scales assess a variety of factors with regards to stress and coping strategies, such as smoking and consuming alcoholic beverages, some of which may not be considered socially acceptable. Therefore, participants may underreport socially undesirable attitudes and behaviours while over-reporting more desirable ones, leading to inaccurate results (Latkin et al., 2017).

### Implications and Areas for Future Research

This study highlights the need for further in-depth understanding of sources of stress, and its impact on medical students’ academic achievement and wellbeing. This will assist in developing awareness and support programs accordingly. Further understanding can also assist in exploring organizational, interpersonal, and personal contributing factors. Additionally, this can contribute towards promoting healthy coping strategies and a supportive environment.

To gain a more thorough understanding of the global impact of stress on medical and non-medical individuals, future studies should aim to assess stress levels and coping strategies among various medical institutions and countries for a more effective and accurate generalisability. To further increase the validity of the results, one can ensure a more parallel age distribution between the two groups to ensure that age does not play a factor in the level of stress experienced by an individual. Conclusively, a larger sample size in future research will allow for further descriptive statistics and inferential statistics.

## CONCLUSION

Stress has had a profound impact globally and this study has attempted to shed light on the level of stress among medical students, i.e. our future physicians, in Dubai, UAE. The results of this study highlighted the higher rates of perceived stress among medical students in comparison to non-medical participants. On the other hand, analysis revealed medium to high rate of engaging in healthy coping strategies among medical students, i.e. mean score of 21.7 out of 32 for problem-focused coping. An individual’s coping response is a key indicator of their psychological adjustment and wellbeing (Heffer and Willoughby, 2017), making it imperative for increased awareness on the matter to protect the mental and physical health of individuals and ensure a healthy adulthood and learning environment for prosperous growth.

## Data Availability

All data produced in the present work are contained in the manuscript

## ACKNOWLEDGEMENTS

We would like to express our deep gratitude to all the participants in the study for their cooperation. Special thanks to Dr. Tom Loney, Associate Professor of Public Health and Epidemiology at MBRU, Dr. Aida J. Azar, Associate Professor of Epidemiology at MBRU, and Dr. Amar H. Khamis, Associate Professor of Biostatistics at MBRU, for their guidance and support. We thank Ms. Vineeta Mohan, Senior Executive, and Mr. Ahmad Al Awadhi, Senior Director of Students Services and Registration at MBRU, for their support. We also thank Mr. Justin Galloway, Senior Mental Health Nurse at Al Jalila Children’s Specialty Hospital, for reviewing the manuscript.

## SETTING

The medical students who have participated in this study are enrolled in Mohammed Bin Rashid University of Medicine and Health Sciences (MBRU) in Dubai, United Arab Emirates.

## ETHICAL APPROVAL

No individuals were excluded from this study based on racial, gender, religious or cultural backgrounds. Consent was obtained from participants age 18 years and above. Assent was obtained from participants below age 18 years along with consent from their parents. This study was conducted with the approval of the local ethical review board at Mohammed Bin Rashid University of Medicine and Health Sciences (Topic Number SRP-2018-040).

## AUTHORS’ CONTRIBUTIONS

Ms. Yas Kaveh Boushehri contributed by conducting the literature search and review, developing the electronic survey, obtaining consent from the participants, study design, data analysis, and manuscript preparation. Prof. Lakshmanan Jeyaseelan contributed to reviewing the data analysis in depth and conducting additional analyses. He also contributed to the manuscript preparation. Dr. Meshal A. Sultan contributed to developing the research question, study design, literature review, revision of the data analysis, manuscript preparation and supervised the research project.

